# Use of home pulse oximetry to enhance remote COVID-19 monitoring: Evaluating a single centre experience

**DOI:** 10.1101/2022.11.07.22281915

**Authors:** Megan W France, Angus H Porter, Cameron J Bennett, Kate L McCarthy

## Abstract

**Introduction:** Telehealth and remote monitoring of patients of patients with mild COVID-19 infection have developed rapidly in response to the pandemic. Many healthcare systems have embraced telehealth for remote clinical monitoring and pulse oximetry for enhanced monitoring.

**Methods:** The experience of a large healthcare centre’s COVID Virtual Ward was reviewed retrospectively with a particular focus on evaluating the effectiveness, safety and utility of finger pulse oximetry within the home. Data from a 2 month period in early 2022 during a BA1 Omicron wave was retrospectively reviewed.

551 high risk patients were issued with pulse oximeters for monitoring oxygen saturations within their home. All patients received daily clinical review via telephone by a nurse or doctor. The group was highly vaccinated with only 6.4% of the cohort unvaccinated. Oximeters were promptly delivered via a courier service across a vast geographical area.

**Results:** Pulse oximetry was well utilised by almost all patients. Only 2.7% of the cohort demonstrated resting oxygen saturations of <94% during their Virtual Ward admission. 91% of patients reporting dyspnoea were able to be safely managed without escalation to an emergency department due to reassuring clinical and oximetry assessment. Home oxygen saturations correlated well with saturations recorded within the ED, with no patients found to have lower saturations compared with home saturations.

**Discussion:** Within a high risk population experiencing COVID-19 infection, pulse oximetry was a useful tool in clinical assessment, correlated well with hospital-based pulse oximetry and allowed a substantial reduction in COVID-19 related ED presentations.

## INTRODUCTION

A cluster of pneumonia cases in Wuhan City, Hubei Province was first reported by Chinese Authorities to the World Health Organisation (WHO) 31 December 2019.^1^ A novel Coronavirus SARS-CoV-2 was subsequently identified as the underlying cause in January 2020.^1^ A rapid increase in international case reports and widespread transmission presented a substantial risk for all healthcare systems. A rapid increase in the number of people requiring care for COVID-19 related disease required health systems to evolve and innovate quickly to meet the unprecedented need for healthcare.^1, 2^

Internationally, care of those with mild COVID-19 related illness within the home via remote models of care has been established as a means of reducing burden upon hospitals and community care. Published accounts of COVID Virtual Care has taken many forms but models usually involve remote monitoring by clinicians utilising phone or video calls into the patient’s home on a regular basis.^1-4^ Some models have focussed upon facilitating early discharge from hospital rather than hospital avoidance.^1,2^ Many models of remote monitoring have also included use of oximetry.^1-8^

Pulse oximetry was first developed in the 1970s for monitoring of oxygen saturations under supervision by a medical practitioner.^3^ Finger pulse oximeters have subsequently evolved towards smaller devices with increasing ease of use, reduced cost and increased accessibility for the general public.^3^

The effectiveness of home oximetry in monitoring people with COVID-19 illness managed at home has been acknowledged to be a potentially useful tool with some potential limitations. Pulse oximeters determine an absorbance ratio and then utilise an internal algorithm to estimate the oxygen saturation.^4, 5^ Saturation measurements below approximately 80% are often unreliable due to data extrapolations utilised to estimate oxygenation.^4-6^ Reduced blood flow through the extremities is a well-documented pitfall of fingertip oximeters, in addition to the potential for poor reading with artificial nails and nail polishes.^4, 5^ Patients with tremor may face challenges with recording accurate measurements.^4, 5^ Additionally, patients with significant dyspnoea and elevated respiratory rate can deceptively have normal oxygen saturations prior to fatigue and respiratory failure^4^. Patients with increased skin pigmentation can register falsely elevated oxygen saturations on finger oximeters, thus potentially missing hypoxaemia.^4, 5, 7-11^

Studies of the reliability of a range of finger oximeters have been published with many outperforming the standard to which they were manufactured.^7, 12^ One such study identified that good sensitivity for hypoxia was achieved with a cut-off value of 94%.^12^ The challenges of accurate oximetry measurements within a moderate to severely unwell cohort of patients with COVID-19 has been documented.^4-6, 12^ We hypothesise that the greatest utility of home oximetry is use in a mild to moderate COVID-19 illness population when accompanied by regular clinical assessment.

This retrospective cohort review of a high risk population with mild to moderate COVID infection aims to

1. Determine the rate of hypoxia identified on home oximetry in a cohort of patients with mild COVID infection.
2. Identify the symptom profile most predictive of hypoxia, radiological abnormality and admission to hospital.
3. Define the role of home oximetry in safely avoiding Emergency Department presentations.

## METHODS

### Model of Care

The Covid Virtual Ward is an initiative designed to ensure safe and effective care of patients recovering from mild to moderate COVID-19 at home.^13^A secondary aim of the model of care is to reduce COVID-19 related presentations to Emergency Departments (ED) within the Metro North Health Service. Metro North Health is the largest Hospital and Health Service within Queensland, Australia, providing care for a population approaching 900 000 living within a 4000 square km geographical area. ^14, 15^ The Virtual Ward also provided care to patients within two other health services within Queensland.

The service commenced in March 2020 and has provided care to 3127 patients during the study period (1^st^ February 2022 to 31^st^ March 2022). ^16^ The COVID-19 Virtual Ward is a large multidisciplinary team comprised of medical practitioners (Specialist, General Practitioner and Resident Medical Officers), nursing staff, pharmacists and administration officers. Clinical services are provided across a 13 hour period, 7 days per week and include an on-call service for advice in the event of patient self-reported deterioration.^13^

Patients were referred to the COVID Virtual Ward with several potential pathways including, self-registering a RAT (Rapid Antigen Test), opting into the service via a positive PCR (Polymerase Chain Reaction), referral from a General Practitioner, referral from an ED or referral from a hospital specialty service.

Oximetry was offered to patients with the following risk factors:

- >65 years old (yo)
- >50yo person identifying as an Aboriginal or Torres Strait Islander person.
- >50yo with co-morbidities including immunosuppression, diabetes mellitus requiring therapy, obesity (Body Mass Index >30), chronic renal impairment, cardiovascular disease (including hypertension), chronic respiratory disease.
- Pregnancy-Trimester 2 and 3.

COVID-specific therapies utilised during the study period included Budesonide and Sotrovimab. Molnupirivir, Nirmatrelvir/Ritonavir started to become available in a limited capacity during the study period. The BA.1/BA.1.1 Omicron strains were predominant during the study period with BA.2 emerging during March 2022.^17^

Oximeters were delivered to a patient’s home via a courier service that operated 7 days per week to enable a same day or next day service. Patients were advised to record their oxygen saturations three times daily and results were reviewed daily by a clinician. At admission to the Virtual Ward, patients were advised to advise the team if any saturations below 94% at rest were recorded.

The cost of providing home oximetry comprises the purchase and delivery of the item to the patient’s home. The purchase of each pulse oximeter (inhealth Pulse Oximeter model A310000) was $27.11. Delivery cost on weekdays ranged from $30 to $160, dependent upon the distance involved. Cost of delivery on weekends involved a $100 surcharge in addition to the baseline delivery cost. To ensure cost-effectiveness, COVID-19 therapies (if criteria for prescription were met) and oximeters for each patient were delivered during the same courier visit.

Assessment of risk of severe COVID-19 related illness was determined by use of an ATLAS (Admission Triage and Level of Care Assignment System) Tool and telephone calls made daily to review symptomatology and oximetry results.^13^ A score was recorded based upon a daily assessment tool (COMPASS) to provide a quantitative assessment of daily symptoms.^13^Qualitative assessment and the COMPASS were noted within an electronic medical record (Virtual Care System platform), viewable by Virtual Ward staff, Health Service Staff and General Practitioners within primary care.

### Data collection

Oximetry data was verbally reported by patients to the clinician providing clinical review that day.

Pharmacy records were reviewed to identify those patients issued with a home oximeter. The VCS was then reviewed for relevant clinical data. Clinical notes and investigations performed within Metro North Health Hospitals was accessed via The Viewer.

This study was assessed by the Metro North Human Research Ethics Committee as a negligible risk research and provided with an exemption from ethics review (EX/2022/QRB/85043).

## RESULTS

### Population

A total of 375 females (including 88 pregnant females) and 176 males were provided with oximeters to support in-home monitoring between 1 February 2022 and 31 March 2022. The median age was 52 years (range: 7-93yo) with a median LOS (Length of Stay) of 6 days (range: 1-24 days).

The study group was highly vaccinated with 328 (59.6%) patients having received 3 or 4 vaccinations and 178 (32.2%) having received 2 vaccinations. Only 35 patients (6.4%) within the cohort were unvaccinated.

There was an elevated level of co-morbidity within in the group, indicated by 460 (83.5%) patients being assigned a very high (VH)- or high (H)-risk level.

### Utilisation of oximetry

Resting oximetry was well utilised and documented for most patients (94.9%). Documentation of walking oximetry was infrequent, with only 31 patients (5.6%) having this parameter noted within their clinical record. Only 15 patients (2.7% of the total oximetry cohort) demonstrated resting home oxygen saturations <94% during their Virtual Ward admission.

### Validation of oximetry readings

Eight patients within the desaturation cohort were escalated for ED review. Of the 7 patients not escalated to ED there were 2 who refused, 3 were determined to have stable, chronically low saturations and 2 who had recent ED review. Five of the desaturation cohort had chronic lung disease including asthma and pulmonary malignancy.

Home oxygen saturations were found to correlate well with saturations recorded upon review within the ED. Oxygen saturations recorded within the ED were found to be the same or higher in all patients referred for desaturation. Importantly, no patients were found to have lower saturations than had been recorded on the home pulse oximeter.

### Respiratory Symptoms and oximetry

Of the patients monitored via oximetry, dyspnoea was frequently reported, with 232 (42.3%) of patients describing dyspnoea at some point during their COVID illness. Table 1 and 2 outline the respiratory symptomatology within the cohorts.

**Table 1:** Frequency of respiratory symptoms during COVID 19 illness in total oximetry cohort.

| **Respiratory Symptom** | **Number of Patients (%)** |
| --- | --- |
| Cough | 515 (93.4%) |
| Dyspnoea | 232 (42.1%) |
| Chest Pain | 54 (9.8%) |
| Chest Tightness | 41 (7.4%) |
| Nil Respiratory Symptoms | 29 (5.2%) |

**Table 2:** Symptoms with those recording desaturation.

| **Symptom** | **Number** |
| --- | --- |
| SOB | 12 |
| CP | 4 |
| Tightness | 4 |
| Cough | 15 |

SOB (Shortness of breath/dyspnoea), CP (chest pain).

Dyspnoea was reported in 12 of the 15 patients (80%) who desaturated. There were 220 patients of the non-desaturation oximetry cohort who reported dyspnoea and were not escalated due to a lack of hypoxia identified on home oximetry.

### Desaturation Cohort

The 15 patients who desaturated had a lower median age of 40 years (24-93 yo) but with a similar predominance of females (67%) compared with the overall oximetry cohort (68%). The median age of the total cohort was 50yo. Dyspnoea was more frequently reported in those patients who desaturated. Most of the desaturation cohort had received 3 or 4 vaccinations against COVID-19 (67%) compared with 59% of the oximetry cohort. Isolated cough was infrequently associated with desaturation with only 2 patients with isolated cough recording oxygen saturations below 94%.

### Escalation to Emergency Department

A total of 45 patients were referred to an Emergency Department (ED) for in-person review (8.2%) due to a clinical concern that could not be resolved by virtual assessment and not due to low oximetry. An additional 4 patients underwent Queensland Ambulance Service (QAS) without transfer onto an ED due to reassuring clinical assessment and clinical measures.

### Radiology and oximetry

An abnormal chest x-ray was described in 16 patients of the total oximetry cohort escalated for in person clinical review. Most of these patients (75%) did not have evidence of desaturation at rest on home testing, reflecting mild viral pneumonitis. Shortness of breath was the symptom that best predicted abnormal radiology, with 15 of the 16 patients describing dyspnoea before their ED attendance.

### Desaturation Cohort

Thoracic imaging was performed in most of the desaturation cohort who presented for ED review. Eleven patients had a chest x-ray (CXR) performed with 8 noted to be normal and 3 abnormal, displaying evidence of acute infiltrates consistent with pneumonitis. Two patients from the normal CXR group proceeded to have a computerised tomography pulmonary angiography (CTPA) and both studies were normal with no evidence of pulmonary embolism or pneumonitis. Only 1 patient required admission due to severe viral pneumonitis.

## DISCUSSION

Our study describes real life validation of home pulse oximetry and daily virtual clinical review in a population of highly vaccinated, high-risk patients with mild to moderate COVID-19 infection. A small minority of patients demonstrated acute hypoxia at rest, indicating that even high-risk patients can be safely supported to recover from COVID-19 infection at home.

Home finger pulse oximetry was well utilised by patients and staff with results documented in the clinical notes of most patients. Assessment of patient related experience measures demonstrated over 99% of patients reported a positive experience with their Virtual Ward admission^18^. This high level of patient acceptance has been previously reported by other studies who have described excellent patient satisfaction with virtual care during the COVID-19 pandemic.^2, 19-23^

Aside from the near ubiquitous symptom of cough, dyspnoea was the symptom most frequently identified within the hypoxic group and thus patients reporting dyspnoea as a prominent symptom of their COVID-19 symptom profile should have access to home pulse oximetry. Conversely, cough in the absence of other respiratory symptoms was not a useful predictor of clinical deterioration, hypoxia and escalation to an ED. This finding may allow further rationalisation of our use of home oximetry towards those who report dyspnoea as a symptom of their COVID-19 illness.

Only a small proportion of the total oximetry-monitored population required escalation to an ED and admission was documented in less than a third of this group. Our low rate of ED escalation of 8.2% was lower than has been reported previously by some groups. Additionally, our rate of home measured hypoxia was very low at 2.7%. A US study reported an ED attendance rate of COVID-19 positive patients of 33% with 20% of the whole cohort developing hypoxia.^19^ A Swedish study published in 2020 reported 25% of their cohort demonstrated hypoxia.^2^ Our low rates of hypoxaemia and escalation are likely contributed to significantly by a high level of vaccination and the generally milder acute clinical course with the BA-2 variant that was predominant during the study period.

The ability to distinguish those with hypoxia from normoxia within a breathless group is invaluable for reducing ED presentations, whilst ensuring clinical safety within the home environment. Our study identified more than a third of patients (38.5%) reporting dyspnoea who avoided escalation to an ED due to reassuring home oxygen saturation measurements alongside a reassuring clinical assessment, even within a high-risk cohort. This has clear benefits for reducing burden upon the healthcare system during COVID-19 waves. Studies performed earlier in the pandemic have also confirmed that patient measured home oximetry safely reduces Emergency Department attendance.^2, 24^ Decreased mortality has also been documented within a high risk, co-morbid South African population.^25^

Our model of care utilised oxygen saturations of 94% for identifying patients potentially requiring escalation for in-person review at an Emergency Department. This level was chosen to reduce the risk of unidentified hypoxaemia and to account for the margin of error with home oximetry devices.^8, 26^ Previous studies have utilised a lower saturation threshold of 92% and confirmed that 92% is a safe threshold for identifying those patients requiring admission for deterioration.^2^ Good correlation between home oxygen saturations and ED measured saturations was found. Reassuringly, our home oximeters did not overestimate oxygen saturations. A multi-centre Australian and New Zealand study established good correlation between oxygen saturations measured via pulse oximeters and PaO2 as measured via arterial blood gas.^27^ This study established that SaO2 of greater than or equal to 92% indicates that hypoxia is not present.^27^ Oxygen saturations of <92% were found to have 95% sensitivity and 90% specificity for PaO2 <60mmHg.^27^ Our use of oxygen saturations of 94% as an alert for further clinical review is safe and unlikely to result in overlooking significant hypoxia.

Our Virtual Ward service plans to alter our assessment of oxygenation to include an assessment of walking oximetry for those with oxygen saturations at or below 94% to further optimise assessment.

Dyspnoea, isolated or in conjunction with other respiratory symptoms, was the symptom most frequently identified amongst those patients who were escalated to ED and displayed abnormal radiology. However, most patients with an acutely abnormal CXR did not show evidence of desaturation on home monitoring. This highlights the occurrence silent viral pneumonitis in the context of COVID-19. Measurement of walking oximetry may have improved the predictive value of abnormal radiology but was not widely recorded in our cohort. This will become an important future focus of our home oximetry monitoring protocols.

The results of this study can guide safe and cost-effective use of oximetry for patients being supported during in-home COVID-19 recovery. Home oximetry measurements should especially be targeted towards high-risk groups who report dyspnoea associated with their COVID infection. Isolated cough, in the absence of a very high-risk profile, is associated with a low risk of hypoxia, escalation to ED and abnormal radiology. The absence of any respiratory symptoms in our study, was with a very low risk of hypoxia or escalation to an ED.

The use of daily clinician review alongside home pulse oximetry allowed opportunities for optimal clinical monitoring, education re COVID-19 infection, COVID-19 recovery, public health guidance and support for patients during isolation.

Monitoring via home oximetry, when combined with care delivered via a virtual team, can reduce emergency department presentations and enhance the safety of care within the home. Our findings allow the targeting of home oximetry monitoring towards the highest risk groups.

## Data Availability

All data produced in the present study are available upon reasonable request to the authors.

## ACKNOWLEDGEMENTS

The authors gratefully acknowledge the staff of the Metro North Health COVID-19 Virtual Ward for their dedication to clinical care throughout the pandemic.

## REFERENCES

1. WHO. WHO Timeline-COVID-19, https://www.who.int (2020, accessed 02.09.22 2022).

2. Shah S, Majmudar K, Stein A, et al. Novel Use of Home Pulse Oximetry Monitoring in COVID-19 Patients Discharged From the Emergency Department Identifies Need for Hospitalization. Acad Emerg Med 2020; 27: 681–692. 2020/08/12. DOI: 10.1111/acem.14053.

3. Michard F, Shelley K and L’Her E. COVID-19: Pulse oximeters in the spotlight. J Clin Monit Comput 2021; 35: 11–14. 2020/06/25. DOI: 10.1007/s10877-020-00550-7.

4. Luks AM and Swenson ER. Pulse Oximetry for Monitoring Patients with COVID-19 at Home. Potential Pitfalls and Practical Guidance. Ann Am Thorac Soc 2020; 17: 1040–1046. 2020/06/11. DOI: 10.1513/AnnalsATS.202005-418FR.

5. Lipnick MS, Feiner JR, Au P, et al. The Accuracy of 6 Inexpensive Pulse Oximeters Not Cleared by the Food and Drug Administration: The Possible Global Public Health Implications. Anesth Analg 2016; 123: 338–345. 2016/04/19. DOI: 10.1213/ANE.0000000000001300.

6. Gurun Kaya A, Oz M, Akdemir Kalkan I, et al. Is pulse oximeter a reliable tool for non-critically ill patients with COVID-19? Int J Clin Pract 2021; 75: e14983. 2021/10/13. DOI: 10.1111/ijcp.14983.

7. Stell D, Noble JJ, Kay RH, et al. Exploring the impact of pulse oximeter selection within the COVID-19 home-use pulse oximetry pathways. BMJ Open Respir Res 2022; 9 2022/02/11. DOI: 10.1136/bmjresp-2021-001159.

8. Administration TG. Limitations of pulse oximeters and the effect of skin pigmentation-medical device safety update https://www.tgs.gov.au/publication-issue/limitations-pulse-oximteres-and-effect-skin-pigmentation (2022), accessed 02.09.22 2022).

9. Valbuena VSM, Barbaro RP, Claar D, et al. Racial Bias in Pulse Oximetry Measurement Among Patients About to Undergo Extracorporeal Membrane Oxygenation in 2019-2020: A Retrospective Cohort Study. Chest 2022; 161: 971–978. 2021/10/01. DOI: 10.1016/j.chest.2021.09.025.

10. Sjoding MW, Dickson RP, Iwashyna TJ, et al. Racial Bias in Pulse Oximetry Measurement. N Engl J Med 2020; 383: 2477–2478. 2020/12/17. DOI: 10.1056/NEJMc2029240.

11. Bickler PE, Feiner JR and Severinghaus JW. Effects of skin pigmentation on pulse oximeter accuracy at low saturation. Anesthesiology 2005; 102: 715–719. 2005/03/26. DOI: 10.1097/00000542-200504000-00004.

12. Schrading WA, McCafferty B, Grove J, et al. Portable, consumer-grade pulse oximeters are accurate for home and medical use: Implications for use in the COVID-19 pandemic and other resource-limited environments. J Am Coll Emerg Physicians Open 2020; 1: 1450–1458. 2021/01/05. DOI: 10.1002/emp2.12292.

13. Ferry OR, Moloney EC, Spratt OT, et al. A Virtual Ward Model of Care for Patients With COVID-19: Retrospective Single-Center Clinical Study. J Med Internet Res 2021; 23: e25518. 2021/02/03. DOI: 10.2196/25518.

14. Metro North Health-About us, https://www.metronorth.health.qld.gov.au/about-us (2022, accessed 02.09.22 2022).

15. Metro North Hospital and Health Service Annual Report 2020-2021, https://www.metronorthhealth.qld.gov.au/about-us/publications/annual-report (2020-21, accessed 02.09.2022 2022).

16. Mackay I. Who requires admission from a Virtual Ward to hospital care in the setting of a COVID-19 Omicron BA.1 and BA.2 outbreak? Manuscript in preparation, Metro North Health, Manuscript in preparation, 2022.

17. Health Q. SARS-CoV-2 Genetics Report. 2022.

18. Report MNH-CaOH. Virtual Ward PREMS. June and July 2022 2022. Metro North Health.

19. Gootenberg DB, Kurtzman N, O’Mara T, et al. Developing a pulse oximetry home monitoring protocol for patients suspected with COVID-19 after emergency department discharge. BMJ Health Care Inform 2021; 28 2021/07/25. DOI: 10.1136/bmjhci-2021-100330.

20. Vaughan L, Eggert LE, Jonas A, et al. Use of home pulse oximetry with daily short message service messages for monitoring outpatients with COVID-19: The patient’s experience. Digit Health 2021; 7: 20552076211067651. 2021/12/21. DOI: 10.1177/20552076211067651.

21. Daly B, Lauria TS, Holland JC, et al. Oncology Patients’ Perspectives on Remote Patient Monitoring for COVID-19. JCO Oncol Pract 2021; 17: e1278–e1285. 2021/06/05. DOI: 10.1200/OP.21.00269.

22. Grutters LA, Majoor KI, Pol-Mattern Esk, et al. Home-monitoring reduces hospital stay for COVID-19 patients. Eur Respir J 2021; 58 2021/09/26. DOI: 10.1183/13993003.01871-2021.

23. Isautier JM, Copp T, Ayre J, et al. People’s Experiences and Satisfaction With Telehealth During the COVID-19 Pandemic in Australia: Cross-Sectional Survey Study. J Med Internet Res 2020; 22: e24531. 2020/11/07. DOI: 10.2196/24531.

24. Dirikgil E, Roos R, Groeneveld GH, et al. Home monitoring reduced short stay admissions in suspected COVID-19 patients: COVID-box project. Eur Respir J 2021; 58 021/04/03. DOI: 10.1183/13993003.00636-2021.

25. Nematswerani N, Collie S, Chen T, et al. The impact of routine pulse oximetry use on outcomes in COVID-19-infected patients at increased risk of severe disease: A retrospective cohort analysis. S Afr Med J 2021; 111: 950–956. 2021/12/25. DOI: 10.7196/SAMJ.2021.v111i10.15880.

26. Administration UFaD. Pulse Oximeter Accuracy and Limitations : FDA Safety Communication, https://www.fda.gov/medical-devices/safety-communications (2021, accessed 31.08.2022 2022).

27. Pilcher J, Ploen L, McKinstry S, et al. A multicentre prospective observational study comparing arterial blood gas values to those obtained by pulse oximeters used in adult patients attending Australian and New Zealand hospitals. BMC Pulm Med 2020; 20: 7. 2020/01/11. DOI: 10.1186/s12890-019-1007-3.

